# Epidemiological Patterns and Characteristics of Animal Bite Cases in Sylhet, Bangladesh: A Retrospective Study of 6,565 Cases

**DOI:** 10.64898/2026.04.21.26351359

**Authors:** Hemayet Hossain, A.S.M. Mohiuddin, Saiful Islam, Khadiza Akter Brishty, Sojib Ahmed, Md. Insan, Masud Parvej, Shyam Kumar Yadav, Shihab Ahmed, Sarna Rani Das, Md. Mahfujur Rahman, Md. Masudur Rahman, Bashudeb Paul

## Abstract

**Background:** Animal bites represent a significant public health concern due to the risk of injuries and transmission of zoonotic diseases such as Rabies, particularly in low and lower- middle-income countries (LMICs). Understanding the epidemiological characteristics of animal bite incidents is essential for improving the prevention and control strategies. This study aimed to characterize the epidemiological patterns and characteristics of animal bite cases in Sylhet, Bangladesh.

**Methodology/Principal findings:** We conducted a retrospective analysis of 6,565 animal bite cases reported between January 1 and December 31, 2024, in Sylhet, Bangladesh. Data on demographic characteristics, type of biting animal, site of bite, and exposure category were collected and analyzed to determine associations using correlation analyses and chi-square tests. Among the victims, 3,917 (60%) were male and 2,648 (40%) were female and young adults aged 20–39 years comprised the largest group (39% of cases). The majority of cases (88.1%) originated from urban areas within Sylhet City Corporation. Cats were the leading cause of bites (56.6%), followed by dogs (35.0%) and monkeys (7.5%), suggesting a shift from the traditional dog-dominated pattern. The most frequently affected anatomical sites were the legs (50.3%) and hands (40.9%). Most exposures were classified as World Health Organization (WHO) Category II (98.2%). Bite incidents showed moderate seasonal variation, with peaks in spring and early autumn. A significant declining temporal trend was observed for monkey bites (R = −0.59, *p* = 0.044), whereas cat and dog bite patterns remained relatively stable throughout the year. Significant associations were identified between bite site and age group, as well as between biting animal and demographic characteristics (*p* < 0.05).

**Conclusion/Significance:** These findings highlight the epidemiological patterns of animal bites in Sylhet and emphasize the need for strengthened public awareness, surveillance, and preventive strategies to reduce animal bite incidents and associated zoonotic disease risks.

**Synnopsis:**

- A large-scale retrospective analysis of 6,565 animal bite cases revealed a cat-dominant bite pattern (56.6%), contrasting with the traditional dog-dominant paradigm in South Asia.
- Young adults (20–39 years) and males (60%) were disproportionately affected, reflecting occupational and behavioral exposure risks.
- Urban residents (88.1%) accounted for the majority of cases, highlighting the growing public health burden of animal bites in rapidly urbanizing settings.
- The most frequently affected anatomical sites were the legs (50.3%) and hands (40.9%). Bite incidents showed moderate seasonal variation, with peaks in spring and early autumn.
- Category II exposures (98.2%) predominated, indicating a high burden of seemingly minor injuries that may be underestimated in rabies prevention strategies.

## Introduction

Every year, over 29 million people require hospital or clinic visits for post-exposure treatment [1]. Most of these cases occur in low-income countries where unvaccinated stray animal population is high, with inadequate post-exposure prophylaxis facilities [2]. Skin breaches caused by animal bites can facilitate the transmission of several zoonotic diseases among which rabies is the most fatal zoonosis [3]. The etiological agent is the rabies virus (RABV), an RNA virus belonging to the *Lyssavirus* genus within the family *Rhabdoviridae* [4]. After entering the human body, the virus initially replicates in myocytes and then enters the neuromuscular junction. It subsequently travels toward the central nervous system (CNS) via retrograde axonal flow along peripheral nerves. Following replication in the CNS the virus disseminates throughout the body [5]. All mammals can act as a host of rabies. Dogs, foxes, and wolves are the main reservoir hosts, whereas cats and monkeys typically act as incidental hosts [6]. Clinically, rabies presents mainly in two forms – furious and paralytic and is almost invariably fatal once clinical signs appear [6].

Around 59,000 deaths from rabies are recorded globally each year. The vast majority (around 95%) occur across Asia and Africa [7]. Children account for nearly around 40% of all victims. Dog bites are responsible for approximately 99% of human rabies deaths although cats, monkeys, and other wildlife also contribute to transmission within the community [1]. Moreover, this disease causes approximately USD 8.6 billion lost every year [7], which is why rabies is classified as a priority Neglected Tropical Disease [1].

This problem is particularly common in South Asia, where the bite incident is higher due to several reasons such as unplanned urban growth, dense human communities, and large numbers of free-roaming animals [8]. In Bangladesh, this remains a significant public health concern, with an estimated 1.6 million free-roaming dogs [9]. For a long time, these animals were the main driver of rabies cases nationwide. Vaccination programs have led to gradual decline in dog-mediated cases [10]. At the same time, less visible shift is also occurring. Rising household incomes and rapid urban growth have together produced a substantial growth in domestic pet ownership. The pet care market is currently growing at approximately 20% per year and is valued at around USD 40-50 million, with cats being the most commonly adopted species [11–13]. As more animals enter households, a proportion of bite incidents are moving from street settings into domestic environments. Surveillance systems here were designed primarily for dog bite cases and were never properly updated for other animals [9]. Monkeys are another emerging concern as they move freely in many districts, bite people, and can also transmit rabies [14–15].

There are significant gaps in the available research. Most of the research consists of dog bite studies in rural settings [10, 16], while research conducted in urban areas remains limited. In addition, most studies have not reported adequate information about the demographics of the victims (i.e., their age, gender, etc.) nor have they identified the species responsible for the bites.

Additionally, the type of injury sustained is not frequently reported in the majority of studies. Documentation of cat bites and monkey bites in official records is extremely rare. As a result, insufficient data are available for policymakers to make informed decisions on developing effective interventions [16–17]. In particular, lack of detailed information on bite severity, the exposure category, and urban populations that may be at risk, the true situation has not been represented in the current literature and thereby limiting rabies control efforts.

The Sylhet City Corporation exhibits a true human-animal interface characterized by pet ownership, numerous strays, and an abundance of monkeys [15]. Due to the nature of the human-animal interface, many people come into the hospital for treatment of bites daily. In this study, 6,565 suspected bite cases were collected from patients attending at Shaheed Shamsuddin Ahmed Hospital in Sylhet during the 1^st^ January to 31^st^ December, 2024. This study aimed to determine the distribution of species responsible for bites, the demographics of the victims, temporal distribution, the anatomic location of the bites on the victims, and the categories of exposure experienced.

## Methods

### Ethical Consent

Ethical approval for this study was obtained from the Institutional Ethics Committee of Teesta University (Approval No: *TU/IEC/2025/006*). The study was conducted in accordance with relevant ethical guidelines for research involving human participants. This study also holds permission letter from Shaheed Shamsuddin Ahmed Hospital, Sylhet. As this study used retrospective, anonymized secondary data, the requirement for informed consent was waived. All data were de-identified prior to analysis to ensure confidentiality.

### Study Design and Setting

A retrospective, hospital-based observational study was conducted to investigate the epidemiological patterns and determinants of animal bite cases in Sylhet, Bangladesh **(Fig 1)**. The study utilized routinely collected clinical records from the Anti-Rabies Vaccination (ARV) unit of Shaheed Shamsuddin Ahmed Hospital, a major tertiary care referral center serving Sylhet City Corporation and surrounding regions. The study period spanned from January 2024 to December 2024.

**Fig 1.**
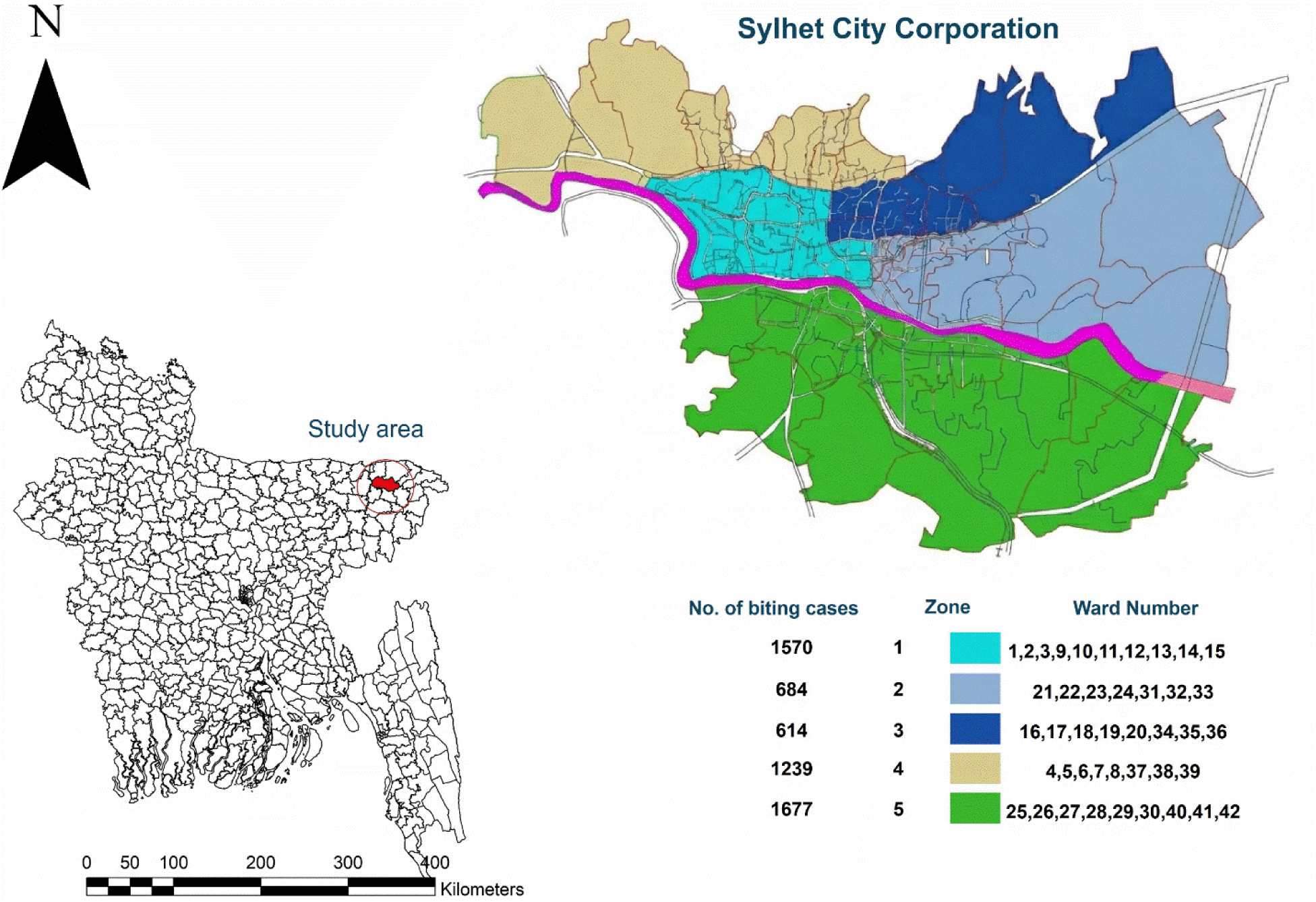
Study area map showing the Sylhet City Corporation (SCC) area along with number of biting cases on city corporation area. The primary shape file for the base layer of the map was extracted from GADM (https://gadm.org/) and the map was generated by ArcGIS software (ArcMap 10.8).

### Study Population and Sample Size

All registered cases of animal bite victims who presented to the ARV unit during the study period were eligible for inclusion. A total of 6,565 cases were included using a census sampling approach, ensuring comprehensive coverage of all reported incidents. Cases with incomplete or missing key epidemiological variables (e.g., age, sex, animal type, or bite site) were excluded (n = 22) to maintain data integrity.

### Data Collection and Variables

Data were extracted from hospital records using a structured data extraction form. Variables collected included:

- **Demographic characteristics:** age, sex, and residence (inside vs. outside city corporation)
- **Exposure characteristics:** type of biting animal (e.g., cat, dog, monkey, fox, others)
- **Clinical characteristics:** anatomical site of bite (head/neck, face, trunk, hands, legs, multiple sites)
- **Exposure severity:** categorized according to World Health Organization (WHO) guidelines (Category I, II, III)

All data were anonymized prior to analysis to ensure patient confidentiality.

### Operational Definitions

Animal bite exposure categories were defined according to WHO standard:

- **Category I:** touching or feeding animals, licks on intact skin
- **Category II:** minor scratches or abrasions without bleeding
- **Category III:** single or multiple transdermal bites or scratches, contamination of mucous membranes

Age groups were stratified into seven categories (0–1, 1–5, 6–12, 13–19, 20–39, 40–64, ≥65 years) to facilitate epidemiological comparisons. To capture finer temporal variation, the study period was categorized into six traditional Bangladeshi seasons (“Shad Ritu”), each comprising two months.

### Statistical Analysis

Data were entered, cleaned, and analyzed using statistical software (e.g., R version 4.5.1). Descriptive statistics were computed, including frequencies, percentages, and distributions of categorical variables.

Bivariate analyses were performed to assess associations between:

- Bite site and demographic variables (age, sex)
- Bite site and animal type
- Animal type and demographic characteristics
- Bite category and exposure characteristics

The Pearson’s Chi-square (χ²) test was used to evaluate statistical significance for categorical variables. To further interpret significant χ² associations, standardized residuals (SRs) were calculated. A *p-value* < 0.05 was considered statistically significant.

## Results

### Demographic Characteristics of Animal Bite Victims

A total of 6,565 animal bite victims were recorded during the study period. Among the victims, 3,917 (60%) were male and 2,648 (40%) were female, indicating a higher occurrence of animal bite injuries among males (**Table 1**).

**Table 1.** Demographic characteristics of animal bite victims.

| Characteristics | Number of Victims<br>N = 6,565 | Percentage<br>(%) |
| --- | --- | --- |
| **Sex** |  |  |
| Male | 3,917 | 60 |
| Female | 2,648 | 40 |
| **Age groups (Years)** |  |  |
| Infants (0-1) | 49 | 0.7 |
| Toddlers (1-5) | 568 | 8.7 |
| Children (6-12) | 1,013 | 15 |
| Teenagers (13-19) | 840 | 13 |
| Young adults (20-39) | 2,582 | 39 |
| Adults (40-64) | 1,314 | 20 |
| Older adults ($\geq 65$) | 199 | 3.0 |
| **Residence** |  |  |
| Inside the City Corporation | 5,784 | 88 |
| Outside of City Corporation | 781 | 12 |

The age distribution showed that young adults aged 20–39 years constituted the largest proportion of cases with 2,582 (39%), followed by adults aged 40–64 years with 1,314 (20%). Children aged 6–12 years accounted for 1,013 (15%), while teenagers aged 13–19 years represented 840 (13%) of the cases. Younger children aged 1–5 years comprised 568 (8.7%), whereas infants aged 0–1 year represented 49 (0.7%). The lowest proportion of cases was observed among older adults aged ≥65 years with 199 (3.0%).

Regarding residence, the majority of the victims were from inside the Sylhet City Corporation area with 5,784 (88%) cases, while 781 (12%) cases were reported from outside the city corporation.

### Characteristics of Animal Bites

A total of 6,565 animal bite cases were recorded, of which 5,784 (88.1%) occurred within the Sylhet City Corporation (SCC) area (Table 2). Cats were the leading cause of bites (56.6%), followed by dogs (35.0%) and monkeys (7.5%) (**Fig 2**) and this distribution remained constant within the SCC (Table 2). The majority of bites occurred on the lower extremities (50.3%) and hands (40.9%), while injuries to the face, trunk, and head/neck were comparatively less frequent. Regarding exposure severity, most cases were classified as WHO Category II (98.2%), with only a small proportion categorized as Category I (1.2%) and Category III (0.6%). A similar distribution pattern was observed among cases within SCC.

**Fig 2.**
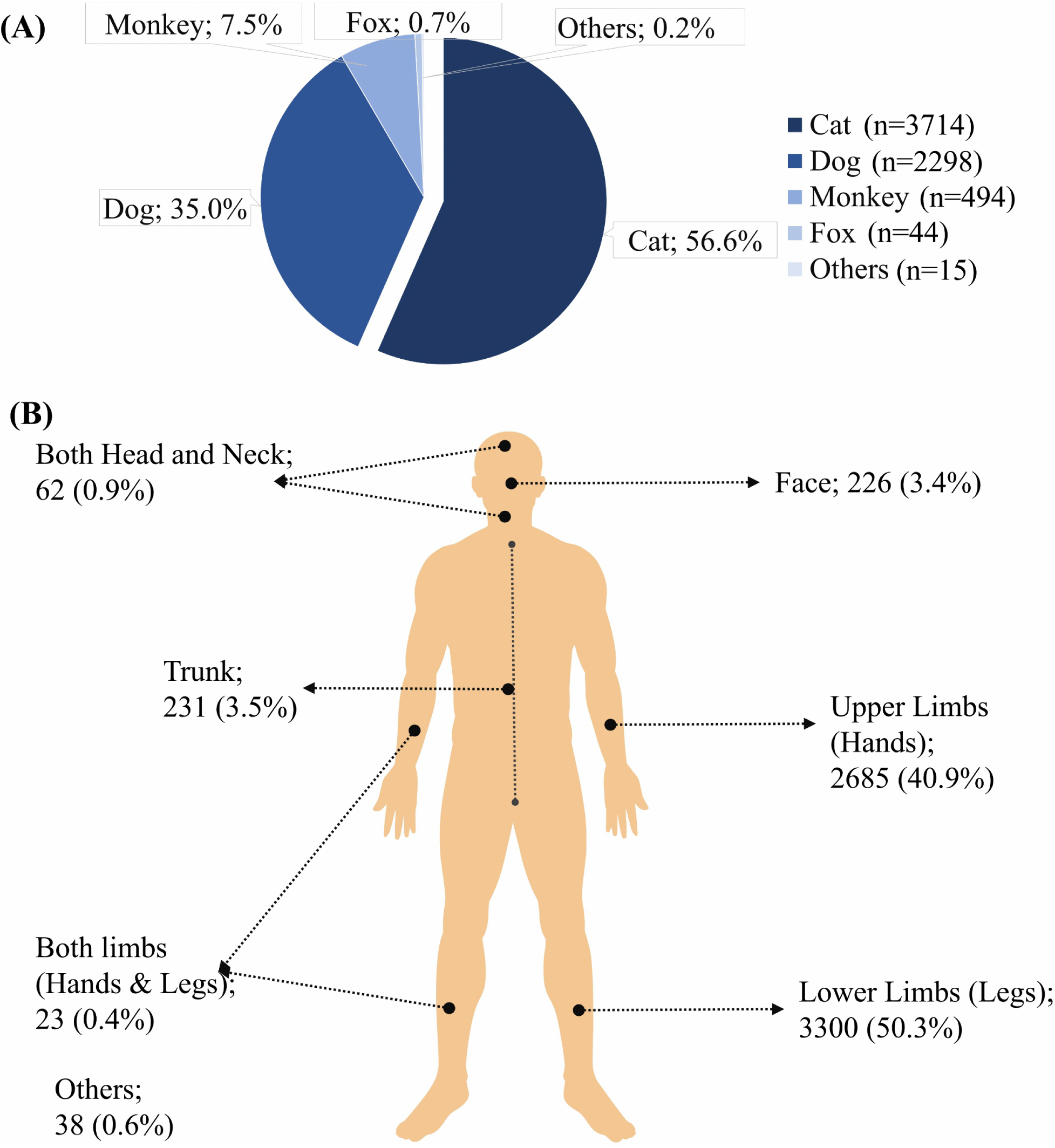
Distribution of animal bite cases and anatomical sites of injury among humans. (A) Proportion of animal species responsible for bite incidents. (B) Anatomical distribution of bite sites among affected individuals.

**Table 2.** Characteristics of animal bites within the Sylhet, Bangladesh.

| Characteristics | Overall Biting Cases |  | Cases within SCC |  |
| --- | --- | --- | --- | --- |
|  | No. of Victims<br>(N = 6,565) | Percentage<br>(%) | No. of Victims<br>(N = 5,784) | Percentage<br>(%) |
| **Biting Animal** |  |  |  |  |
| Cat | 3,714 | 56.6 | 3,314 | 57.3 |
| Dog | 2,298 | 35.0 | 1,968 | 34.0 |
| Monkey | 494 | 7.5 | 464 | 8.0 |
| Fox | 44 | 0.7 | 26 | 0.5 |
| Others | 15 | 0.2 | 12 | 0.2 |
| **Site of Bite** |  |  |  |  |
| Head and Neck | 62 | 0.9 | 53 | 0.9 |
| Face | 226 | 3.4 | 198 | 3.4 |
| Trunk | 231 | 3.5 | 192 | 3.3 |
| Hands | 2,685 | 40.9 | 2,404 | 42.0 |
| Legs | 3,300 | 50.3 | 2,885 | 50 |
| Both hands and legs | 23 | 0.4 | 19 | 0.3 |
| Others | 38 | 0.6 | 33 | 0.6 |
| **WHO Animal Bite Category** |  |  |  |  |
| I | 83 | 1.2 | 71 | 1.2 |
| II | 6,441 | 98.2 | 5,681 | 98 |
| III | 41 | 0.6 | 32 | 0.6 |

### Temporal trends of Animal Bite Cases

Figure 3 shows heterogeneity in the temporal distribution of bite cases by animal source. Cat bites were the most frequent exposure throughout the year, with monthly counts ranging from 70 to 445, followed by dog bites ranging from 33 to 345. In contrast, monkey bites occurred at substantially lower levels (9 to 82 cases/month), while fox and other-animal bites were rare across all months. For dog bites, the monthly pattern showed a modest overall decline over time (R = −0.36, *p* = 0.25), with relatively high counts in January (275) and June (345), followed by lower counts toward July (66) and December (65). Cat bites displayed a similar but non-significant downward trend (R = −0.30, *p* = 0.34), remaining consistently high across most months, with notable peaks in September (445) and June (418).

**Fig 3.**
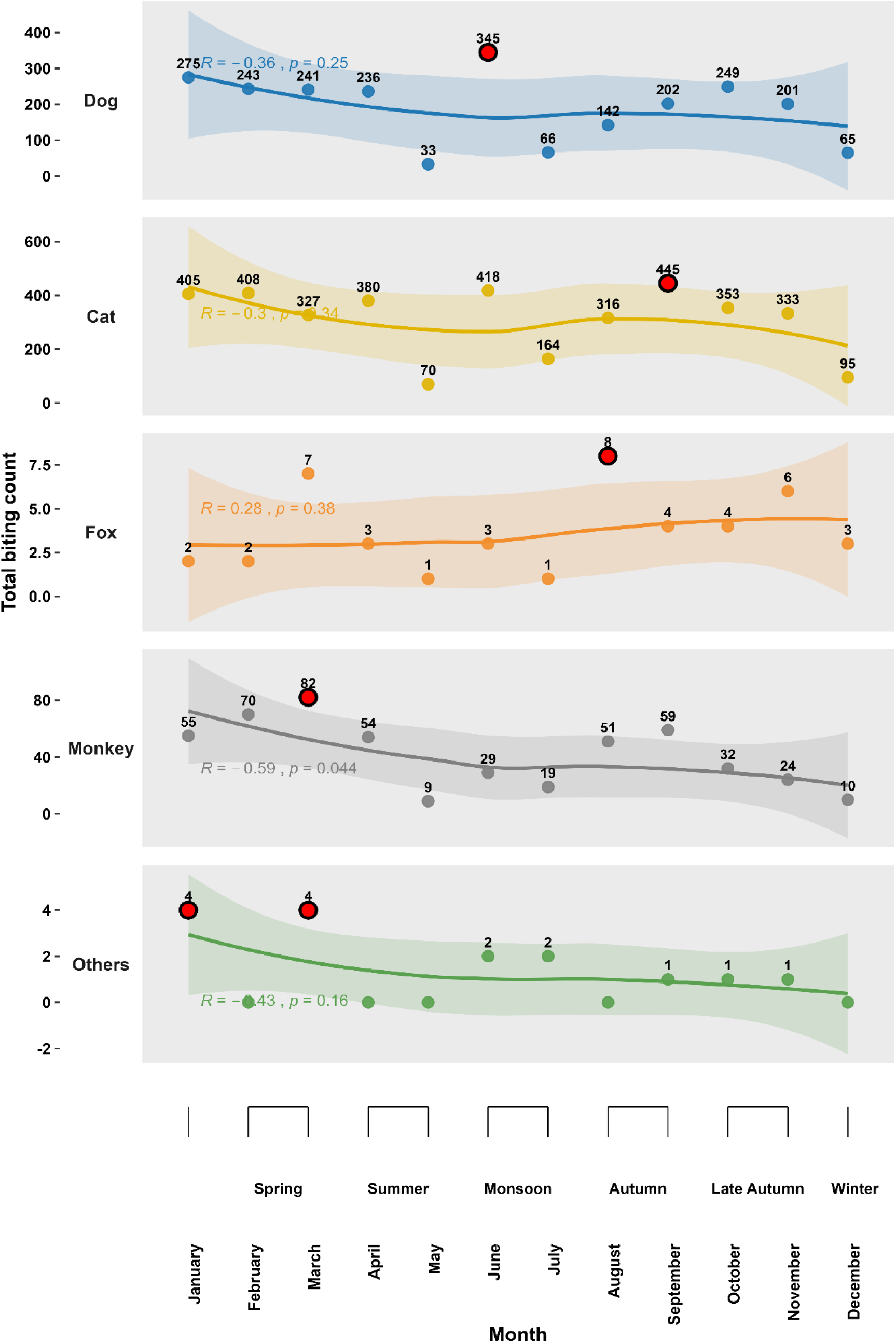
Monthly and seasonal variation in animal bite counts by source animal in Sylhet, Bangladesh. Points show monthly case counts for dog, cat, fox, monkey, and other animals, with smoothing lines and 95% confidence intervals indicating temporal trends. Correlation coefficients (R) and *p*-values are shown for each animal group. Red circle indicates highest biting value in month.

By contrast, monkey bites showed a statistically significant declining temporal pattern (R = - 0.59, *p* = 0.044). Counts decreased from 55–82 cases in the earlier months to 10 cases in December. This was the only animal category demonstrating evidence of a significant monotonic temporal trend.

Across all seasons, dog bites were predominantly localized to the leg, with the highest counts observed in spring (n = 341), late autumn (n = 337), and autumn (n = 271) (Fig 4). Cat bites were concentrated in the hand and leg, particularly hand bites in spring (n = 416) and leg bites in autumn (n = 363). Monkey bites occurred less frequently and involved mainly the hand and leg, with the highest hand-bite count in spring (n = 71). Fox and other animal bites were rare across all anatomical sites and seasons. Overall, the most frequently affected sites were the leg (n = 3,300) and hand (n = 2,685).

**Fig 4.**
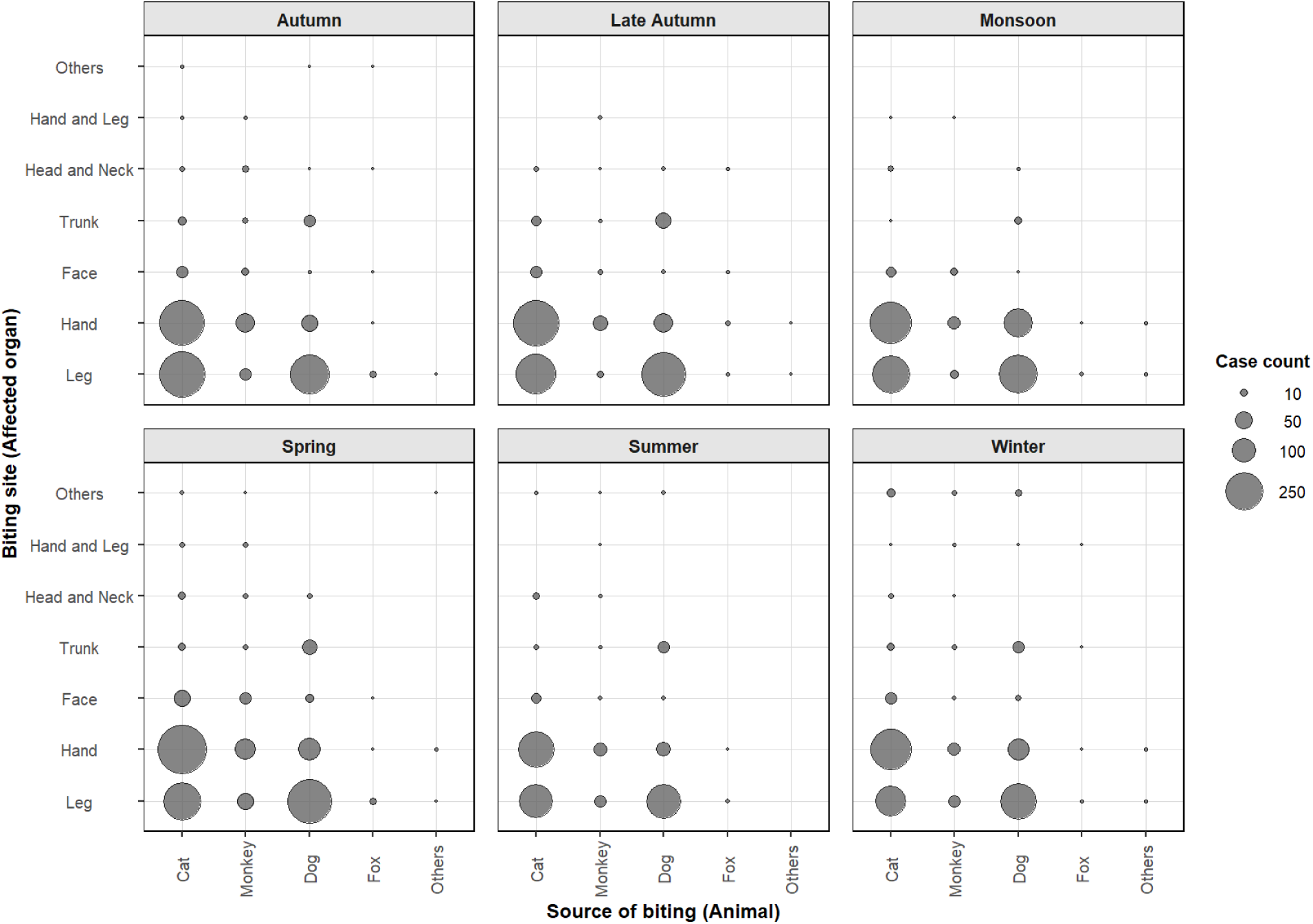
Seasonal distribution of animal bite cases by source animal and affected anatomical site in Sylhet, Bangladesh. Bubble size represents case count for each combination of biting animal and bite site within season-specific panels.

Despite moderate seasonal variation in case volume, the anatomical distribution remained stable, characterized by lower-limb dog bites and hand- and leg-dominant cat bites throughout the year.

### Relationship between Site of Bite with Sex and Age

A statistically significant association was observed between age group and site of bite (*p* < 0.0001) in SCC area. Leg bites were the most common across all age groups. Among young adults aged 20–39 years, 1,177 (41%) bites occurred on the legs and 1,031 (43%) on the hands (**Table 3**).

**Table 3.**
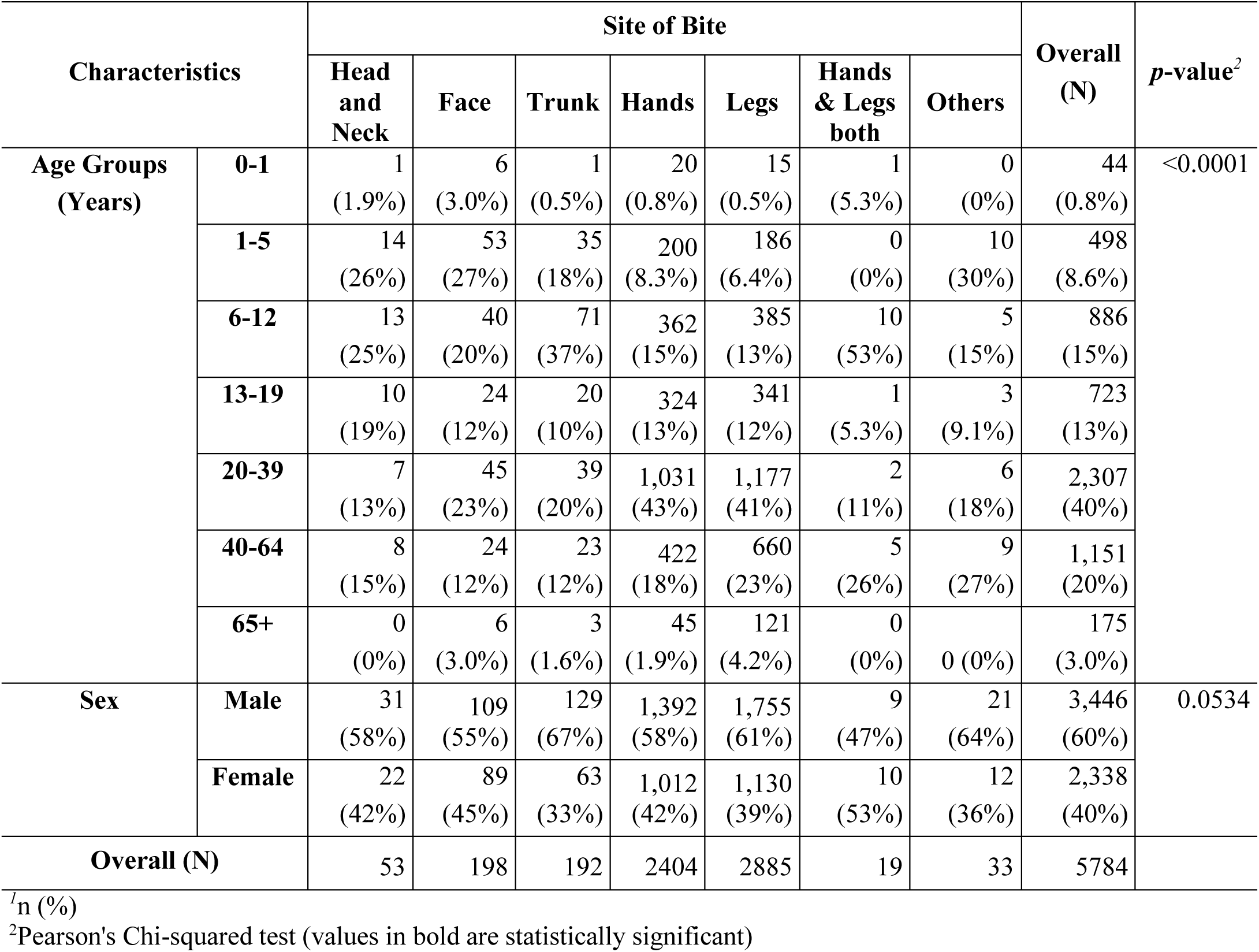
Relationship of the site of bite with sex and age in SCC.

| Characteristics |  | Site of Bite |  |  |  |  |  |  | Overall (N) | *p*-value<sup>2</sup> |
| --- | --- | --- | --- | --- | --- | --- | --- | --- | --- | --- |
|  |  | Head and Neck | Face | Trunk | Hands | Legs | Hands & Legs both | Others |  |  |
| **Age Groups (Years)** | **0-1** | 1<br>(1.9%) | 6<br>(3.0%) | 1<br>(0.5%) | 20<br>(0.8%) | 15<br>(0.5%) | 1<br>(5.3%) | 0<br>(0%) | 44<br>(0.8%) | <0.0001 |
|  | **1-5** | 14<br>(26%) | 53<br>(27%) | 35<br>(18%) | 200<br>(8.3%) | 186<br>(6.4%) | 0<br>(0%) | 10<br>(30%) | 498<br>(8.6%) |  |
|  | **6-12** | 13<br>(25%) | 40<br>(20%) | 71<br>(37%) | 362<br>(15%) | 385<br>(13%) | 10<br>(53%) | 5<br>(15%) | 886<br>(15%) |  |
|  | **13-19** | 10<br>(19%) | 24<br>(12%) | 20<br>(10%) | 324<br>(13%) | 341<br>(12%) | 1<br>(5.3%) | 3<br>(9.1%) | 723<br>(13%) |  |
|  | **20-39** | 7<br>(13%) | 45<br>(23%) | 39<br>(20%) | 1,031<br>(43%) | 1,177<br>(41%) | 2<br>(11%) | 6<br>(18%) | 2,307<br>(40%) |  |
|  | **40-64** | 8<br>(15%) | 24<br>(12%) | 23<br>(12%) | 422<br>(18%) | 660<br>(23%) | 5<br>(26%) | 9<br>(27%) | 1,151<br>(20%) |  |
|  | **65+** | 0<br>(0%) | 6<br>(3.0%) | 3<br>(1.6%) | 45<br>(1.9%) | 121<br>(4.2%) | 0<br>(0%) | 0<br>(0%) | 175<br>(3.0%) |  |
| **Sex** | **Male** | 31<br>(58%) | 109<br>(55%) | 129<br>(67%) | 1,392<br>(58%) | 1,755<br>(61%) | 9<br>(47%) | 21<br>(64%) | 3,446<br>(60%) | 0.0534 |
|  | **Female** | 22<br>(42%) | 89<br>(45%) | 63<br>(33%) | 1,012<br>(42%) | 1,130<br>(39%) | 10<br>(53%) | 12<br>(36%) | 2,338<br>(40%) |  |
| **Overall (N)** |  | 53 | 198 | 192 | 2404 | 2885 | 19 | 33 | 5784 |  |

<sup>1</sup><sub>n</sub> (%)
<sup>2</sup>Pearson's Chi-squared test (values in bold are statistically significant)

Among children aged 6–12 years, 71 (37%) bites were reported on the trunk, while 40 (20%) occurred on the face. In younger children aged 1–5 years, relatively higher proportions of bites were observed on the face with 53 (27%) cases and head and neck with 14 (26%) cases.

When stratified by sex, males experienced a higher proportion of bites in most anatomical regions, including 1,755 (61%) bites on the legs and 1,392 (58%) bites on the hands. Females accounted for 1,130 (39%) leg bites and 1,012 (42%) hand bites. However, the association between sex and site of bite was not statistically significant (*p* = 0.0534).

### Association among the Bite Site, Biting Animals, and Bite Category

The site of bite showed a significant association with the type of biting animal (*p* < 0.001) inside SCC area. Cat bites were predominantly reported on the Hands with 1,760 (73%) cases, whereas dog bites were more frequently observed on the Legs with 1,410 (49%) cases and on the trunk with 127 (66%) cases (**Table 4**). Monkey bites showed a relatively higher proportion of injuries involving both upper and Legs with 13 (68%) cases. A significant association was also found between site of bite and exposure category (*p* < 0.001). The majority of Category II bites occurred on the limbs, particularly the Legs with 2,851 (99%) cases and Hands with 2,355 (98%) cases.

**Table 4.** Relationship of the site of bite with the biting animal and the category of bite in SCC area.

| Characteristic |  | Site of Bite |  |  |  |  |  |  | Overall (N) | p-value<sup>2</sup> |
| --- | --- | --- | --- | --- | --- | --- | --- | --- | --- | --- |
|  |  | Head and Neck | Face | Trunk | Hands | Legs | Hands & Legs both | Others |  |  |
| **Biting Animal** | **Cat** | 28<br>(53%) | 130<br>(66%) | 50<br>(26%) | 1,760<br>(73%) | 1,323<br>(46%) | 6<br>(32%) | 17<br>(52%) | 3,314<br>(57%) | <0.001 |
|  | **Dog** | 8<br>(15%) | 18<br>(9.1%) | 127<br>(66%) | 396<br>(16%) | 1,410<br>(49%) | 0<br>(0%) | 9<br>(27%) | 1,968<br>(34%) |  |
|  | **Fox** | 2<br>(3.8%) | 1<br>(0.5%) | 1<br>(0.5%) | 5<br>(0.2%) | 17<br>(0.6%) | 0 (0%) | 0 (0%) | 26<br>(0.4%) |  |
|  | **Monkey** | 15<br>(28%) | 49<br>(25%) | 14<br>(7.3%) | 236<br>(9.8%) | 131<br>(4.5%) | 13<br>(68%) | 6<br>(18%) | 464<br>(8.0%) |  |
|  | **Others** | 0<br>(0%) | 0<br>(0%) | 0<br>(0%) | 7<br>(0.3%) | 4<br>(0.1%) | 0<br>(0%) | 1<br>(3.0%) | 12<br>(0.2%) |  |
| **Category of Bite** | **I** | 1<br>(1.9%) | 3<br>(1.5%) | 4<br>(2.1%) | 40<br>(1.7%) | 22<br>(0.8%) | 0<br>(0%) | 1<br>(3.0%) | 71<br>(1.2%) | <0.001 |
|  | **II** | 50<br>(94%) | 192<br>(97%) | 184<br>(96%) | 2,355<br>(98%) | 2,851<br>(99%) | 19<br>(100%) | 30<br>(91%) | 5,681<br>(98%) |  |
|  | **III** | 2<br>(3.8%) | 3<br>(1.5%) | 4<br>(2.1%) | 9<br>(0.4%) | 12<br>(0.4%) | 0<br>(0%) | 2<br>(6.1%) | 32<br>(0.6%) |  |
| **Overall (N)** |  | 53 | 198 | 192 | 2404 | 2885 | 19 | 33 | 5784 |  |

<sup>1</sup>n (%)
<sup>2</sup>Pearson's Chi-squared test

The relationship between the biting animal and the bite site exhibited a unique pattern. Most dog bites happened on the lower body parts (legs) and showed a strong positive association with leg injuries (Standardized Residual, SR = 24.94) and a moderate association with the trunk (SR = 10.83) (Fig 5). Cat bites were mostly seen on the hands (SR = 22.29), whereas bites to the legs and trunk were infrequent. But monkey bites showed a different pattern. Significant associations have been demonstrated across upper-body regions, but were less frequently observed in the legs. Fox bites showed moderate associations with the face and head–neck regions, suggesting a preference for upper body sites, although these trends were less pronounced. The category of other animals exhibited no strong deviations except for a slight association with less common bite sites.

**Fig 5.**
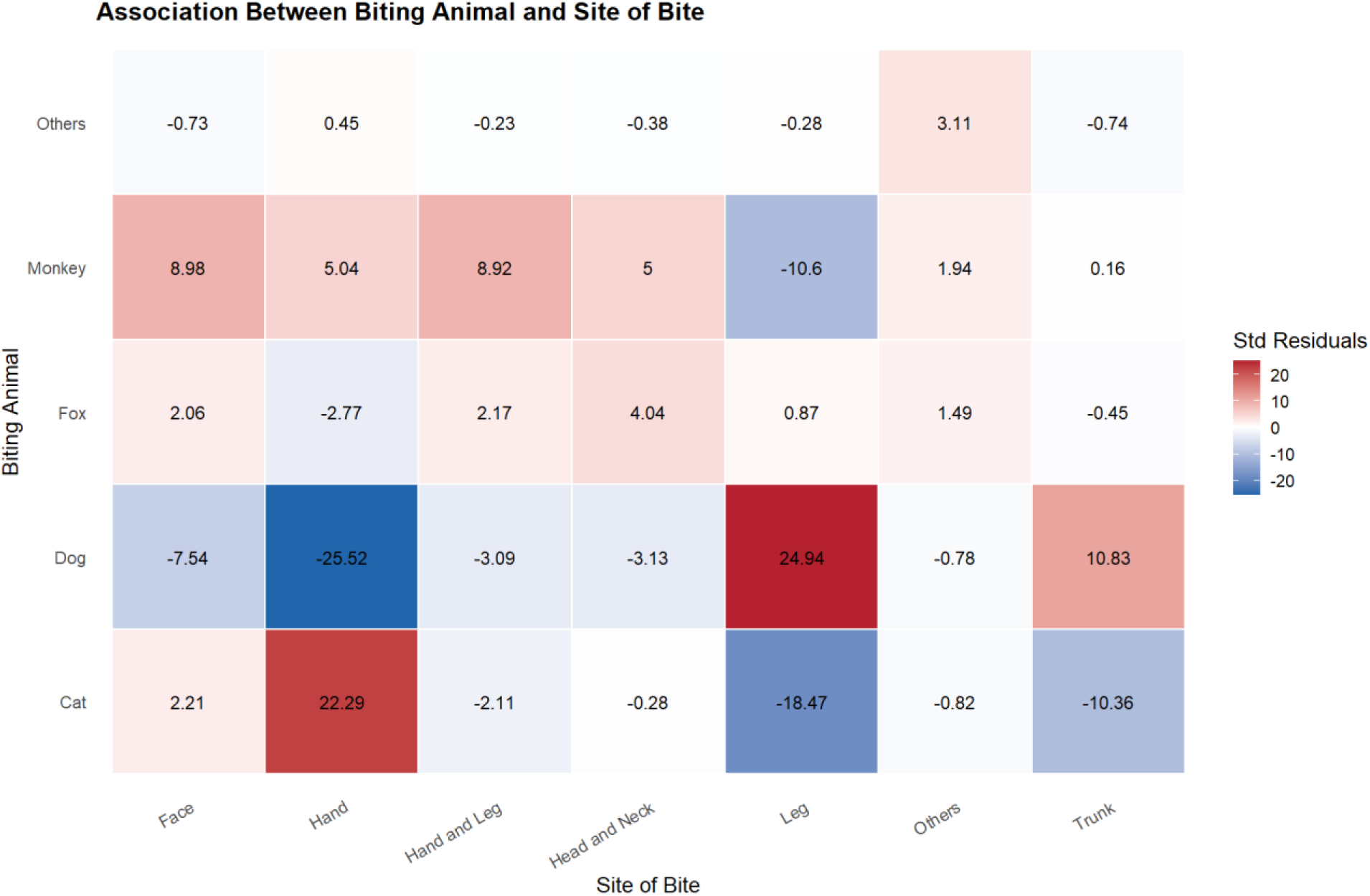
Association between biting animal and anatomical site of bite based on standardized residuals from chi-square analysis. The heatmap illustrates the strength and direction of association between different biting animals (dog, cat, fox, monkey, others) and bite locations (face, hand, hand & leg, head & neck, leg, others, trunk). Color intensity represents standardized residual values: red indicates a higher-than-expected frequency, while blue indicates a lower-than-expected frequency.

### Association between the Biting Animals and Demographic Characteristics

Overall, animal bite cases were concentrated in children, adolescents, and young adults, although age distribution differed across epidemiological groups. By biting category (Fig 6A), category III cases were the youngest (median 12 years), whereas category I and II showed older and broadly similar age distributions (medians 23 and 24 years, respectively). By biting site (Fig 6B), leg bites occurred in relatively older patients (median 27 years), followed by hand bites (median 23 years). In contrast, bites to the trunk, face, and head and neck were concentrated in younger individuals, with median ages ranging from 10 to 13 years. By sex (Fig 6C), females were slightly older than males overall (median 26 vs 23 years). However, both sexes showed wide age dispersion. By animal source (Fig 6D), dog bites involved relatively older patients (median 26 years), while monkey bites were concentrated in younger age groups (median 10 years). Fox and other animal bites also showed comparatively older age profiles (median 27 years each). Temporal variation was modest (Fig 6E). The oldest age profile was observed during the monsoon, particularly in June (median 29 years), whereas May showed the youngest monthly distribution (median 21 years).

**Fig 6.**
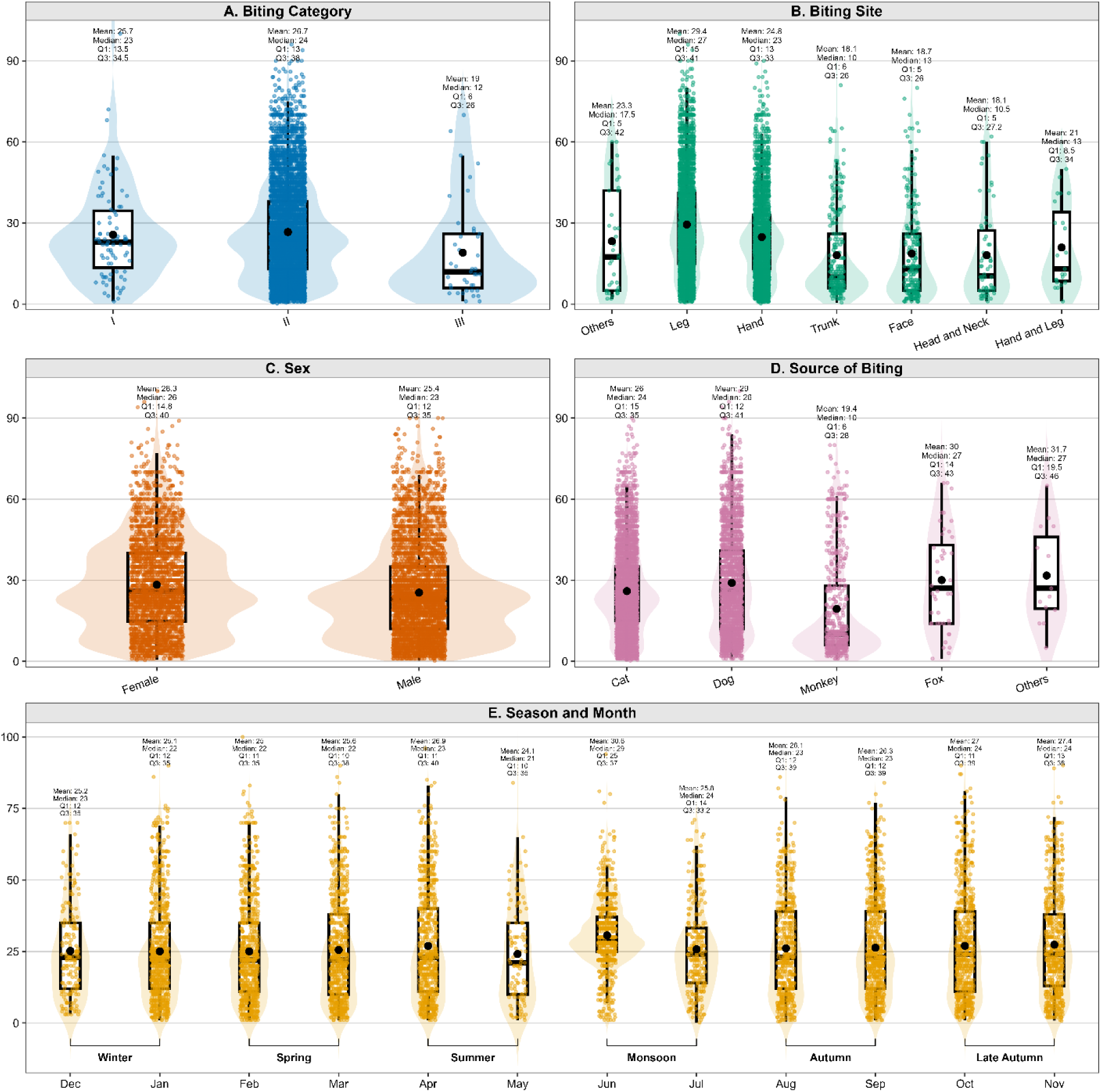
Age distribution of animal bite cases across epidemiological characteristics in Sylhet, Bangladesh. Panels show age by (A) biting category, (B) biting site, (C) sex, (D) source of biting animal, and (E) season and month. Violin plots represent age density, boxplots show median and IQR, black dots indicate mean age, and jittered points represent individual cases.

Overall, younger ages were especially prominent in category III bites, craniofacial/truncal bites, and monkey-related injuries, while relatively older age distributions were observed for dog bites, leg bites, and monsoon-period cases.

A statistically significant association was observed between type of biting animal and sex (*p* < 0.001). Males accounted for 1,771 (53%) cat bite cases and 1,389 (71%) dog bite cases, whereas females represented 1,543 (47%) cat bite cases and 579 (29%) dog bite cases (**Table 5**). Age group was also significantly associated with the type of biting animal (*p* < 0.001). Cat bites were most common among young adults aged 20–39 years with 1,505 (45%) cases, while monkey bites were more frequently observed among children aged 6–12 years with 161 (35%) cases and children aged 1–5 years with 105 (23%) cases.

**Table 5.** Relationship of biting animals with sex, age group and category of bite in SCC area.

| Characteristic |  | Biting Animals |  |  |  |  | Overall (N) | p-value<sup>2</sup> |
| --- | --- | --- | --- | --- | --- | --- | --- | --- |
|  |  | Cat | Dog | Fox | Monkey | Others |  |  |
| Sex | Male | 1,771<br>(53%) | 1,389<br>(71%) | 18<br>(69%) | 260<br>(56%) | 8<br>(67%) | 3,446<br>(60%) | <0.001 |
|  | Female | 1,543<br>(47%) | 579<br>(29%) | 8<br>(31%) | 204<br>(44%) | 4<br>(33%) | 2,338<br>(40%) |  |
| Age Group<br>(Years) | 0-1 | 33<br>(1.0%) | 6<br>(0.3%) | 1<br>(3.8%) | 4<br>(0.9%) | 0<br>(0%) | 44<br>(0.8%) | <0.001 |
|  | 1-5 | 274<br>(8.3%) | 117<br>(5.9%) | 1<br>(3.8%) | 105<br>(23%) | 1<br>(8.3%) | 498<br>(8.6%) |  |
|  | 6-12 | 375<br>(11%) | 348<br>(18%) | 2<br>(7.7%) | 161<br>(35%) | 0<br>(0%) | 886<br>(15%) |  |
|  | 13-19 | 461<br>(14%) | 213<br>(11%) | 5<br>(19%) | 41<br>(8.8%) | 3<br>(25%) | 723<br>(13%) |  |
|  | 20-39 | 1,505<br>(45%) | 721<br>(37%) | 6<br>(23%) | 70 (15%) | 5 (42%) | 2,307 (40%) |  |
|  | 40-64 | 603<br>(18%) | 465<br>(24%) | 9<br>(35%) | 72<br>(16%) | 2<br>(17%) | 1,151<br>(20%) |  |
|  | 65+ | 63<br>(1.9%) | 98<br>(5.0%) | 2<br>(7.7%) | 11<br>(2.4%) | 1<br>(8.3%) | 175<br>(3.0%) |  |
| Category<br>of bite | I | 46<br>(1.4%) | 20<br>(1.0%) | 0<br>(0%) | 4<br>(0.9%) | 1<br>(8.3%) | 71<br>(1.2%) | <0.001 |
|  | II | 3,262<br>(98%) | 1,932<br>(98%) | 21<br>(81%) | 455<br>(98%) | 11<br>(92%) | 5,681<br>(98%) |  |
|  | III | 6<br>(0.2%) | 16<br>(0.8%) | 5<br>(19%) | 5<br>(1.1%) | 0<br>(0%) | 32<br>(0.6%) |  |
| Overall (N) |  | 3,314 | 1,968 | 26 | 464 | 12 | 5,784 |  |

<sup>1</sup>n (%)
<sup>2</sup>Pearson's Chi-squared test (values in bold are statistically significant)

Regarding bite categories, the majority of cases across all animal types were Category II exposures, including 3,262 (98%) cat bites, 1,932 (98%) dog bites, and 455 (98%) monkey bites. A relatively higher proportion of Category III exposures was observed among fox bites with 5 (19%) cases.

## Discussion

This study provides valuable insights into the epidemiology of 6,565 animal bite cases in Sylhet, Bangladesh, highlighting several important patterns with implications for rabies prevention and urban public health interventions. One of the most notable findings is the predominance of cat bites (56.6%), surpassing dog bites (35.0%), which contrasts with the well-established dog-dominated trend documented across South Asian and global contexts. Dogs account for the majority of bite cases worldwide, including in Bangladesh (76.7% in Dhaka) [18], India (92% in Bhopal, 93.6% in Delhi, and 98% in Kashmir) [19–21], Nepal (74% in Kathmandu) [22], and Thailand (77.8% in Eastern Thailand) [23]. However, a study conducted in Mymensingh, Bangladesh, reported a similar pattern to the present findings, with 52.6% of victims bitten by cats [16]. Likewise, a study from Saudi Arabia reported a higher proportion of cat-related exposures, accounting for 75% of cases [24]. The higher proportion of cat bites in this study may be associated with increasing pet ownership and closer human–animal interactions in urban environments. In recent years, a notable expansion of the pet care sector has been observed in Bangladesh, with cats being among the most commonly adopted companion animals [25]. People often handle cats during feeding and play without adequate caution, which can result in superficial injuries such as minor bites or scratches, especially affecting the hands. This may explain the high frequency of hand injuries, as well as the predominance of WHO Category II exposures observed in this study.

The higher number of male bite victims, particularly from dog reported in this study is consistent with several previous findings from South Asia and the Sub-Saharan Africa. Similar trends have been reported in Nigeria (73.3% in Benue State) [26], India (73.43% in Jabalpur and 81.6% in Mumbai) [27, 28], and Iran (76.3% in Kermanshah) [29]. Greater outdoor exposure as well as frequent encounters with stray or free-roaming animals among males in Bangladeshi context [30], may explain the higher frequency of male bite victims compared to females. However, the data is completely reversed for cat bite cases, where female represented the maximum cases. A study conducted in Hong Kong also found a predominance of cat-related injuries among females [31]. Females own more cats worldwide and spend more time with household pets, which may increase their risk of cat bites compared to dog bites

Young adults (aged 20-39 years) constituted the majority of bite victims; aligning with findings from Nigeria [26], India [27], Germany [32], and Turkey [33]. This higher proportion reflects the greater engagement of young adults in occupational and outdoor activities. Adults aged 40-64 years represented the second-largest group, although there is little information about this group in the existing literature. School-aged children (5-18 years) also constituted a notable portion of cases. Although this percentage was lower than that reported in previous studies from Mymensingh, Bangladesh (25.90%) [16], Bhopal, India (34.58%) [19], and Rajasthan, India (25.35%) [34], children are particularly vulnerable because of their behavioral factors and inadequate knowledge to recognize or avoid dangerous interactions with animals. Monkey bites proved to be a primary threat for schoolchildren aged 6–12 (35% of cases), similar to Odisha, India, where children encounter primates near forest edges [35].

The majority of bite victims in this study were from urban areas, where high population density, inadequate waste management, and frequent human–animal interactions results in increased exposure [9]. A higher proportion bite victims form the urban settings have also been reported in Turkey (73.2%) [36] and Ghana (64.7%) [37]. In contrast, approximately 85.6% of victims were reported from rural areas in Ethiopia [38]. These findings underscore the important role of both the environmental setting and lifestyle factors in determining the risk of animal bite exposure.

The proportion of monkey bites in this present study was considerably higher than that reported in other studies from Bangladesh. Monkey bites accounted for only 0.9% of cases in Dhaka [18], and 1.3% in Mymensingh [16]. In Sylhet, the reasons for the higher proportion of monkey bites may be related to the presence of peri-urban tea gardens, religious practice of feeding monkeys at temples or shrines and existence of open waste dumps [39]. Monkey bites are concerning not only because of the percentage, but also due to the anatomical sites of the injuries. It is reported that monkeys caused an unusually high number of dangerous bites to the head and neck. Injuries to those areas are very risky due to proximity to the brain and cause a lower incubation period of rabies.

In this study, the majority of cases were recorded as Category II, contrasting with findings from cities in India such as Mumbai (78.18%) [28], Delhi (91.0%) [20] and Kashmir (57.6%) [21], where Category III cases predominate. As cats are predominant biting species in this study and typically cause minor scratches rather than deep wounds during playing and feeding, it is perhaps the cause for the dominance of Category II in this study. Although treating Category II wounds results in lower medical costs, they still carry risks, as victims may underestimate the danger of minor wounds and delay vaccination. A study conducted in Kathmandu is somewhat consistent with this study, where 70% cases were also under category II [22]. The proportion of fox bites is low, but the number of fox bites categorized as Category III (19%) reflects the potential role of foxes in the transmission of sylvatic rabies. Though it is rare continuous monitoring of these populations is important to prevent spillover into humans.

A statistically significant association was observed between the age of the victims and the anatomical site of the bite. Children aged 1–5 years are particularly vulnerable to craniofacial injuries, which pose a higher risk of rabies transmission due to their proximity to the CNS. This pattern is consistent globally; for example, in Israel, 49.7% of pediatric animal bite cases requiring hospitalization involved craniofacial injuries [40]. Similar findings of increased craniofacial involvement among children have been reported in India (Kashmir) and Korea [21, 41]. The susceptibility of children may be attributed to their explorative toward animals and their shorter height, which places their head and face closer reach of aggressive animals. In addition, young children lack the physical skills for effective defense, resulting both the possibility and severity of injury. The risk of craniofacial injury also persists among school-aged children throughout development. In contrast, young adults (20–39 years) more commonly sustain bites to the upper and lower extremities, which is consistent with findings from Turkey (38.9% upper and 50.5% lower extremities) [33].

Another significant association was reported between the animal species and location of the biting site. Consistent with global findings, limbs are the common biting sites for the maximum number of cases. Of all types of animal bites, just over half of all cat bites occur on the hand. This finding is also consistent with studies conducted in Germany [32] and China [42]. These injuries actually occur during feeding or interaction involving hands reflecting the close association with the pet. Dog bites, on the other hand, involved trunk and legs. When an animal attacks a person who is standing or walking, it is simply easier to reach the legs than anywhere else. The same scenario has also been documented in several studies, such as in India (45.62% in Rajasthan) [34], Nepal (58% in Kathmandu) [22] and Greece (50.6%) [43]. In contrast, bites to upper extremities are most common in the USA (47.3%) due to intentional handling or playing with animals either pets or stray [44]. Although monkey bites were less frequent in this study, there was a relatively higher involvement of the head and neck; such exposures are clinically important due to proximity of the injury to the brain. This distribution of bite severity creates a pattern of "predictability" for physicians in estimating the risk of an individual when designing prevention strategies.

Temporal analysis showed modest seasonal fluctuations, characterized by mild peaks in spring and early autumn; however, overall trends remained stable year-round. In case of monkey bite, a statistically significant declining trend was observed in the present study. While the underlying reason is unclear based on the available data. This may be due to change in several factors like human activity, environmental conditions, or local wildlife dynamics. Research focusing on ecological or longitudinal studies would be helpful to clarify these relationships.

Strategies for preventing rabies in South Asia have predominantly focused on dog bites. However, this study showed that this model is inadequate as cat bites were predominant here. Although there are fewer monkey bites, they tend to occur on the face, head, and neck, which are higher risk for exposure to rabies in humans. Therefore, dog and cat vaccination programs are warranted. Pet owners need better awareness about animal vaccination and safe handling. Monkey bite prevention also requires attention and should not be neglected. Children and women face different risks from different animals, so awareness campaigns need to speak to them directly, not just the general public.

In total, this study analyzed 6,565 cases of animal bites (multiple species) from January to December 2024, comprising the largest recent data set for this region in urban settings. The study allows for many types of analyses by age, gender, type of animal, and severity of exposure - all of which are essential to better develop prevention strategies. However, the study also has limitations; the study only included patients who visited a hospital or similar facility. Patients treated by traditional healers or who do not seek any form of treatment were excluded from the study population. This is a common problem in South Asian studies and means the real number of bites is probably higher. Also, because population data for the area was not available, it was not possible to calculate precise incidence rates.

### Strengths and Limitations

This study represents one of the largest recent datasets on animal bite epidemiology in Bangladesh, incorporating 6,565 cases over a three-year period. The inclusion of multiple animal species (cats, dogs, monkeys, foxes) provides a more comprehensive understanding beyond dog-centric studies. Detailed stratification by demographics, anatomical bite sites, and WHO exposure categories enhances its epidemiological depth and policy relevance. Additionally, the study captures urban transmission dynamics, which remain underexplored in South Asian contexts.

Given the retrospective design, the study relied on hospital-reported cases and may not capture unreported or community-managed bite incidents. Additionally, the absence of population denominator data precluded the calculation of incidence rates.

## Conclusion

This retrospective study provides a comprehensive overview of animal bite epidemiology in Sylhet, Bangladesh. The findings demonstrate that animal bite incidents are more common among males and predominantly affect individuals in the 20–39 years’ age group, reflecting increased occupational and outdoor exposure. Most victims were residents of the Sylhet City Corporation area, indicating a higher risk in urban environments. Cats and dogs were responsible for the majority of bite incidents, with cats representing the largest proportion of cases. The legs and hands were the most frequently affected anatomical sites, suggesting that many bites occur during routine human–animal interactions. Most cases were classified as Category II exposures, highlighting the importance of timely medical care and appropriate post-exposure management to prevent zoonotic infections, particularly Rabies. Overall, the study underscores the need for improved animal bite surveillance, public awareness programs, responsible pet management, and strengthened rabies prevention strategies in Sylhet. These measures will contribute to reducing the burden of animal bite injuries and minimizing the risk of rabies transmission in the community.

### Recommendation

- Expand rabies prevention strategies beyond dogs to include cats and monkeys, reflecting the evolving epidemiological landscape.
- Implement integrated One Health surveillance systems that capture multi-species bite data and urban transmission dynamics.
- Strengthen public awareness programs, particularly targeting young adults, children, and pet owners, emphasizing the risks of minor (Category II) exposures.
- Promote mass vaccination programs for both domestic and stray animals, especially in urban settings.
- Develop community-based reporting systems to capture underreported bite cases and improve national surveillance accuracy.
- Introduce behavioral and risk-factor studies to better understand human–animal interaction patterns and guide targeted interventions.

## Acknowledgements

The authors would like to acknowledge all the participants who directly or indirectly helped in the data collection process of this study.

## Author contributions

**Conceptualization:** Hemayet Hossain, Bashudeb Paul.

**Data curation:** Hemayet Hossain, Saiful Islam, Md. Insan, Sojib Ahmed, A.S.M. Mohiuddin.

**Formal analysis:** Saiful Islam, Hemayet Hossain.

**Investigation:** Hemayet Hossain, Saiful Islam, Md. Insan, Sojib Ahmed, A.S.M. Mohiuddin, Khadiza Akter Brishty, Masud Parvej, Shihab Ahmed, Sarna Rani Das, Shyam Kumar Yadav

**Methodology:** Hemayet Hossain, Saiful Islam, Md. Insan, A.S.M. Mohiuddin, Masud Parvej, Shihab Ahmed, Khadiza Akter Brishty, Sarna Rani Das, Bashudeb Paul

**Resources:** Hemayet Hossain, Saiful Islam, Khadiza Akter Brishty, Md. Mahfujur Rahman, Md. Masudur Rahman, Bashudeb Paul

**Software:** Hemayet Hossain, Saiful Islam, Bashudeb Paul

**Writing – original draft**: Hemayet Hossain, Sojib Ahmed, Md. Mahfujur Rahman, Md. Masudur Rahman, Bashudeb Paul.

**Writing – review & editing:** Hemayet Hossain, Md. Mahfujur Rahman, Md. Masudur Rahman, Bashudeb Paul

**Project administration:** Bashudeb Paul.

**Validation:** Md. Mahfujur Rahman, Md. Masudur Rahman, Bashudeb Paul.

**Visualization:** Md. Mahfujur Rahman, Md. Masudur Rahman, Bashudeb Paul.

**Supervision:** Md. Mahfujur Rahman, Md. Masudur Rahman, Bashudeb Paul.

## Data availability

The authors confirm that all data underlying the findings are fully available without restriction.

## Funding

The author(s) received no specific funding for this work.

## Competing interests

The authors have declared that no competing interests exist.

## Notes

### Competing Interest Statement

The authors have declared no competing interest.

### Author Declarations

Ethical approval for this study was obtained from the Institutional Ethics Committee of Teesta University (Approval No: TU/IEC/2025/006).

